# Multivariate brain-based dimensions of child psychiatric problems: degrees of generalizability

**DOI:** 10.1101/2023.03.12.23287158

**Authors:** Bing Xu, Lorenza Dall’Aglio, John Flournoy, Gerda Bortsova, Brenden Tervo-Clemmens, Paul Collins, Marleen de Bruijne, Monica Luciana, Andre Marquand, Hao Wang, Henning Tiemeier, Ryan L. Muetzel

## Abstract

Multivariate machine learning techniques are a promising set of tools for identifying complex brain-behavior associations. However, failure to replicate results from these methods across samples has hampered their clinical relevance. This study aimed to delineate dimensions of brain functional connectivity that are associated with child psychiatric symptoms in two large and independent cohorts: the Adolescent Brain Cognitive Development (ABCD) Study and the Generation R Study (total *n*=8,605). Using sparse canonical correlations analysis, we identified three brain-behavior dimensions in ABCD: attention problems, aggression and rule-breaking behaviors, and withdrawn behaviors. Importantly, *out-of-sample* generalizability of these dimensions was consistently observed in ABCD, suggesting robust multivariate brain-behavior associations. Despite this, *out-of-study* generalizability in Generation R was limited. These results highlight that the *degree* of generalizability can vary depending on the external validation methods employed as well as the datasets used, emphasizing that biomarkers will remain elusive until models generalize better in true external settings.

## Introduction

Psychiatric neuroimaging has sought to illuminate the neurobiological underpinnings of psychiatric disorders over the past few decades, providing a unique opportunity to study neurodevelopment during childhood and adolescence, a key risk window for the emergence of mental health problems^1^. One surging area of research, brain-behavior association studies, has been a promising approach to explore individual brain variability that predicts behavioral phenotypes^2–4^. To date, however, rigorously validated and generalizable neurobiological biomarkers that are able to guide clinical practice remain elusive^5–9^. Several features of the literature can explain this empirical reality, such as insufficient statistical power, variability across methodologies, and a heavy reliance on univariate analysis techniques that fail to map the multidimensional neural bases of psychiatric disorders^8, 10^. Inherent heterogeneity and high comorbidity of psychiatric disorders exacerbate the problem, rendering it difficult to isolate the most relevant neural features of interest. This is especially the case for children and adolescents who usually present less clearly defined psychopathology and heterotypic continuity of symptoms and phenotypes^11^.

A potential promising path forward is the application of multivariate machine learning techniques^3, 12^. Multivariate methods can assess the covariation of neural phenotypes, jointly modeling different types of information (e.g., brain and behaviors). They are less hampered by the small effect sizes that univariate analyses of psychiatric neuroimaging studies typically observe^8, 10^, resulting in greater statistical power and the potential for better reproducibility^3^. Moreover, multivariate methods with a data-driven nature can shed light on transdiagnostic brain-behavior associations by identifying coherent and specific brain mechanisms that cut across diagnoses^13–15^, offering the potential for parsing possible sources of comorbidity and heterogeneity.

One widely-used multivariate method in psychiatric neuroimaging is canonical correlation analysis (CCA), a technique that aims to identify the common variation across phenotypes and dissect their complex relationships into a small number of distinct dimensions^4^. Several studies have implemented CCA to depict transdiagnostic brain-behavior dimensions^2, 16^, and the identified brain dimensions could be further used to study potential neurobiologically informed classifications of psychiatric disorders^17^. However, the replicability of these methods has come under heavy scrutiny^18–20^. One of the key elements, which is largely missing from previous work, is robust external validation in a fully independent dataset (i.e., not a hold-out subsample from a single cohort). Though this has been widely implemented in the validation of prediction models in medical research^21, 22^, psychiatric neuroimaging studies have not generally adopted these external validation strategies.

In most existing studies, various forms of cross-validation have been implemented by sampling randomly from a pool of data from a single study. This means the data are often highly homogenous in many respects, including participant sampling and data collection protocols. While this step of within-study internal validation is a reasonable start, understanding the real-world generalizability of a model requires a different dataset that is fundamentally distinct from the data used to train the model. This means the model must be robust to sampling and methodological differences, which is a necessity for population-level model generalizability^12^. Without this crucial step of a proper generalizability test, clinical utility will remain unreachable.

The current study aims to address these gaps by leveraging two large population-based neurodevelopmental cohorts, the Adolescent Brain Cognitive Development (ABCD) Study (*n*=6,529) and the Generation R Study^23, 24^ (*n*=2,076), in order to delineate robust and generalizable multivariate associations between resting-state functional magnetic resonance imaging (rs-fMRI) connectivity and child psychiatric symptoms. As childhood and adolescence are periods of marked brain development^25^ during which psychiatric problems emerge or exacerbate^26^, understanding how neural mechanisms are linked to psychopathology during this time is crucial. Using the ABCD study as the discovery set, we applied sparse CCA (SCCA) under a rigorous multiple hold-out framework^27, 28^ to identify linked brain-behavior dimensions. Importantly, the trained model in ABCD was applied and evaluated in a completely independent, external data set to test the *out-of-study* generalizability of the results. Given the two cohorts utilized in this study represent the largest in-site and multisite studies of neurodevelopment in the world, they are uniquely positioned to conduct such multivariate analyses. We highlight the importance of model generalizability in the context of psychiatric neuroimaging and offer several insights on how to improve generalizability through these techniques.

## Results

### Dimensions of child psychiatric symptoms and functional connectivity

A total of 8,605 rs-fMRI scans from the multi-site ABCD Study (ages 9-to-10 years from 21 study sites) and the single-site Generation R Study (ages 9-to-12 years) were summarized using the 352-region Gordon parcellation^29^. After several important functional MRI covariates were regressed out (e.g., motion, see Methods), functional time courses from the different regions (333 cortical, 19 subcortical) were used to construct connectivity matrices for each individual by correlating the time courses pair-wise across all regions. To safeguard against overfitting, the connectivity matrices underwent dimensionality reduction by principal component analysis (PCA) with a weighting scheme (100 components, see Methods). Eight syndrome scales were used to characterize psychiatric symptoms of children assessed by the parent-report Child Behavioral Checklist (CBCL)^24^(anxious/depressed, withdrawn/depressed, somatic, social, aggressive, rule-breaking, thought, and attention problems). The ABCD data were randomly split into a training set consisting of 18 sites (ABCD_Training_) and a test set consisting of 3 sites (ABCD_Test_). This split procedure was repeated 10 times to reduce sampling bias, resulting in 10 pairs of independent train-test sets. Analyses in ABCD_Training_ and ABCD_Test_ sets were fully separated to prevent data leakage (Figure 1). ABCD_Training_ and ABCD_Test_ sets were comparable on age, sex, race/ethnicity/parental education, and psychiatric symptoms (Table 1).

**Figure 1.**
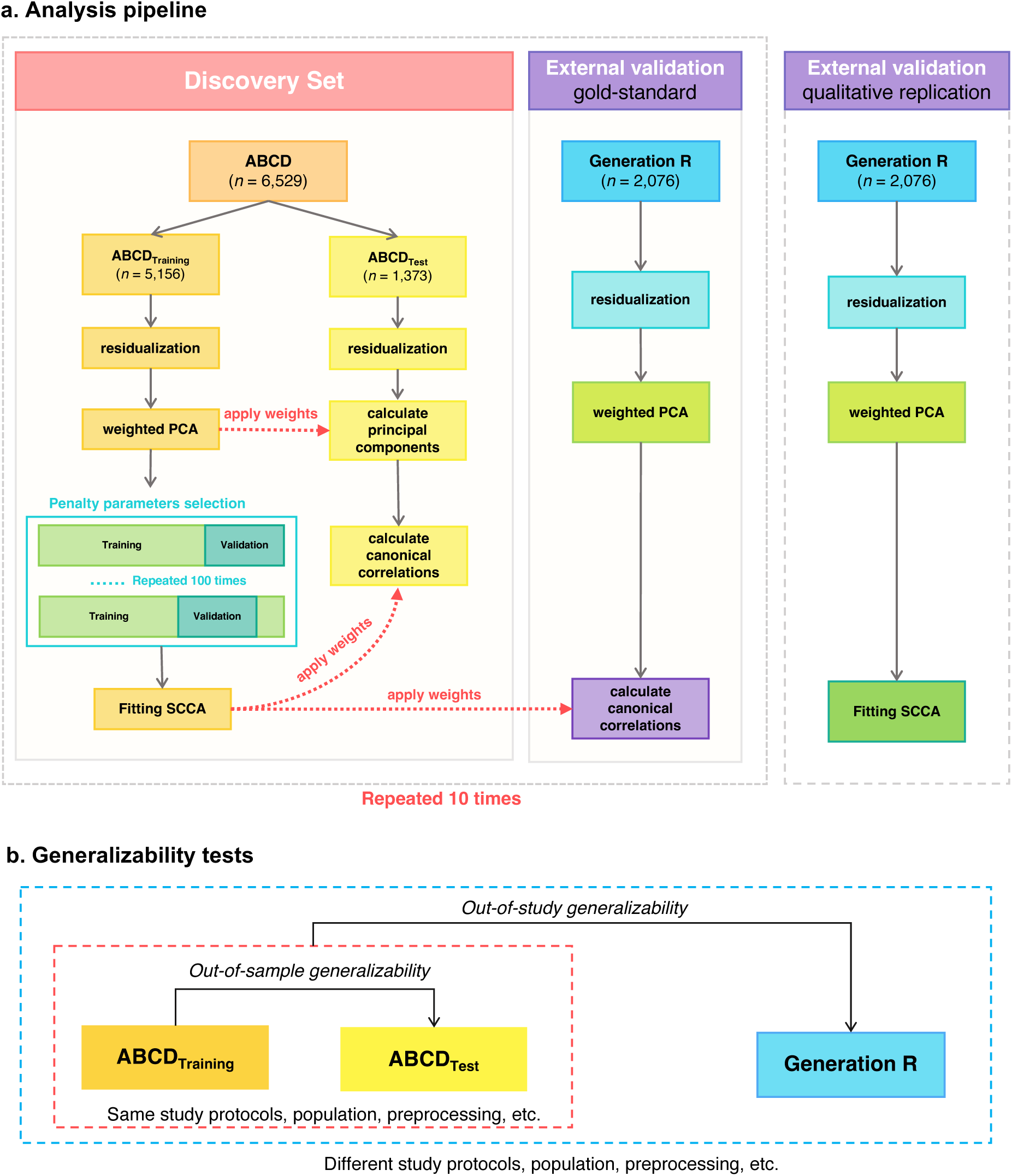
Multivariate brain-behavior associations analysis pipeline. *Note.* **a-b.** ABCD was the discovery set and Generation R as the external validation set. The discovery set was divided into training and test sets 10 times, resulting in 10 train-test pairs in ABCD. The eigenvectors of PCA from the ABCD_Training_ set were applied to ABCD_Test_ set to calculate the principal components, then the weight vectors (canonical loadings) obtained from the ABCD_Training_ set were projected to ABCD_Test_ set to compute the out-of-sample generalizability. Similarly, weight vectors of SCCA from the ABCD_Training_ set were then directly applied to Generation R to assess the out-of-study generalizability of the model. We also implemented the qualitative replication approach, in which we train the CCA model independently in Generation R and compare the results across the two cohorts. Note that the sample size in ABCD is an example from one train-test split.

**Table 1.** Descriptive statistics of the discovery set (example) and the external validation set.

| Discovery set |  |  | External validation set |  |
| --- | --- | --- | --- | --- |
| **ABCD**<br>*n* = 6,529 |  |  | **Generation R**<br>*n* = 2,076 |  |
|  | *ABCD<sub>Training</sub>* | *ABCD<sub>Test</sub>* |  |  |
| N | 5,156 | 1,373 | N | 2,076 |
| Age (years), M(SD) | 10.0 (0.6) | 9.9 (0.6) | Age (years), M(SD) | 9.9 (0.3) |
| Sex |  |  | Sex |  |
| Girls, (%) | 48.9 | 47.2 | Girls, (%) | 52.3 |
| Race/ethnicity (%) |  |  | Nation of birth (%) |  |
| White | 52.9 | 60.1 | Dutch | 65.6 |
| African American | 13.9 | 9.8 | Non-Dutch European | 15.1 |
| Hispanic | 20.7 | 17.5 | Non-European | 19.3 |
| Asian | 2.1 | 1.5 |  |  |
| Others | 10.3 | 10.5 |  |  |
| Parental education (%) |  |  | Maternal education (%) |  |
| Low | 5.5 | 3.9 | Low | 2.9 |
| Medium | 40.5 | 37.3 | Medium | 34.8 |
| High | 54.0 | 58.8 | High | 62.3 |
| Child Behavior Checklist (CBCL) |  |  | Child Behavior Checklist (CBCL) |  |
| Externalizing scores, M(SD) | 4.4 (5.7) | 4.1 (5.9) | Externalizing scores, M(SD) | 3.9 (4.6) |
| Internalizing scores, M(SD) | 5.2 (5.6) | 4.7 (5.4) | Internalizing scores, M(SD) | 4.8 (4.9) |
| Total scores, M(SD) | 18.1 (17.6) | 16.9 (17.8) | Total scores, M(SD) | 17.2 (15.2) |
| Framewise displacement (median) | 0.16 | 0.16 | Framewise displacement (median) | 0.15 |

*Note.* Values are frequencies for categorical variables and means and standard deviations for continuous variables. The descriptive statistics for ABCD were based on one of the ten train-test splits, other splits showed similar statistics. M = Mean, SD = Standard Deviation

### Initial derivation of brain-behavior dimensions

Using the ABCD_Training_ set to train the model, six brain-symptom dimensions (canonical variates) were identified using an elastic net combining LASSO and ridge penalties with SCCA (*r*_1_ = 0.20, *r*_2_ = 0.19, *r*_3_ = 0.17, *r*_4_ = 0.16, *r*_5_ = 0.15, *r*_6_ = 0.13, *p*s < .01 after permutation testing; averaged across 10 splits, see Table 2, Figure 2a).

**Figure 2.**
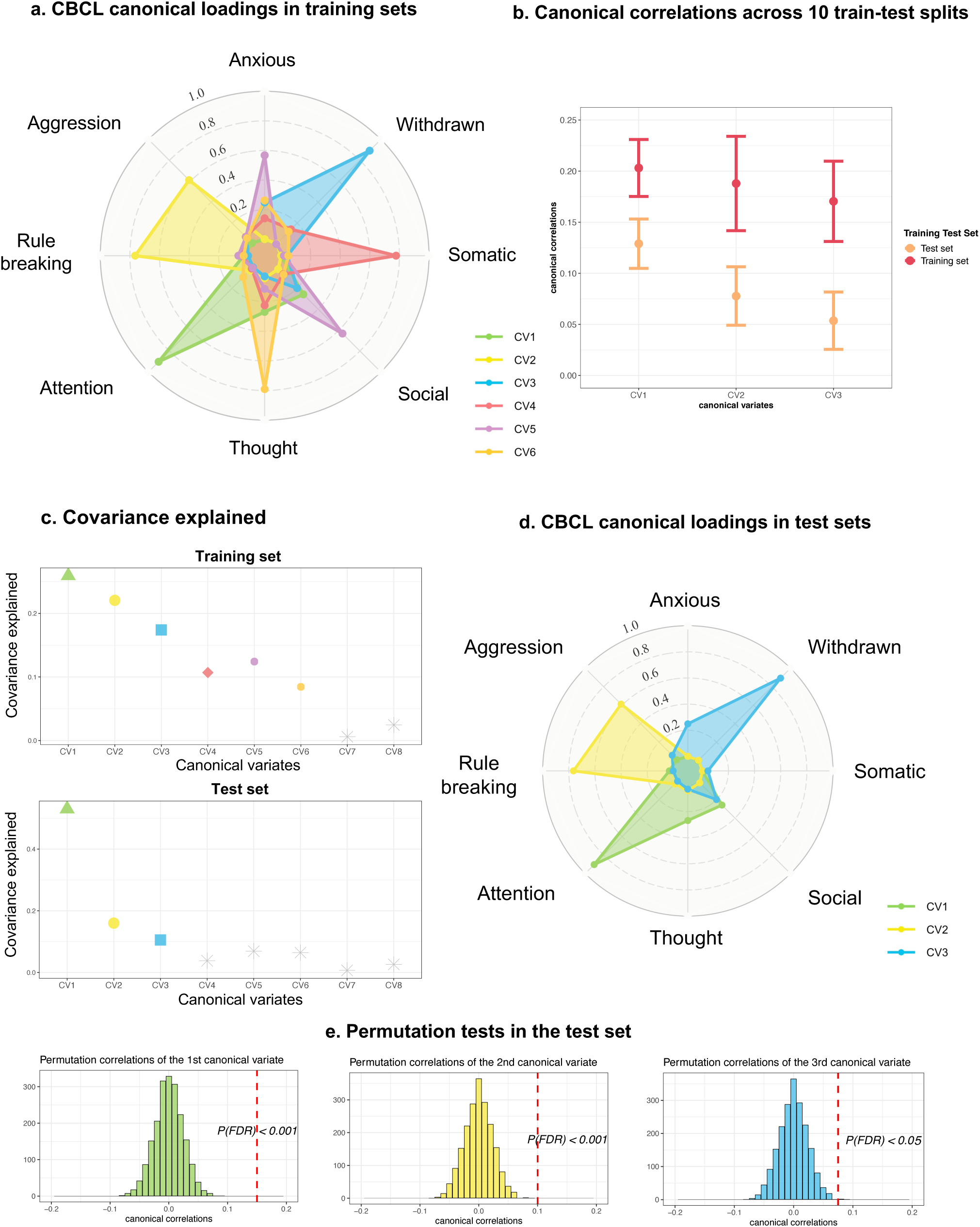
Associated dimensions of brain connectivity and CBCL syndrome scores in ABCD. *Note.* SCCA identified brain-behavior correlates in training and test sets of ABCD. **a.** The first six canonical correlations survived the permutation tests in the ABCD_Training_ sets. The canonical loadings of CBCL syndrome scores in the ABCD_Training_ set were averaged across 10 train-test splits. **b.** The mean and standard deviation of the first three canonical correlations across 10 train-test splits. **c.** Covariance explained in the training and test sets (example from one train-test split). **d.** The first three canonical variates were replicated in ABCD_Test_ set across the 10 train-test splits. **e.** Permutation tests for the first three canonical correlations in the test sets (example from one of the 10 train-test splits), the red dotted lines represent the canonical correlations in the unshuffled data. P values were corrected for multiple testing.

**Table 2.**
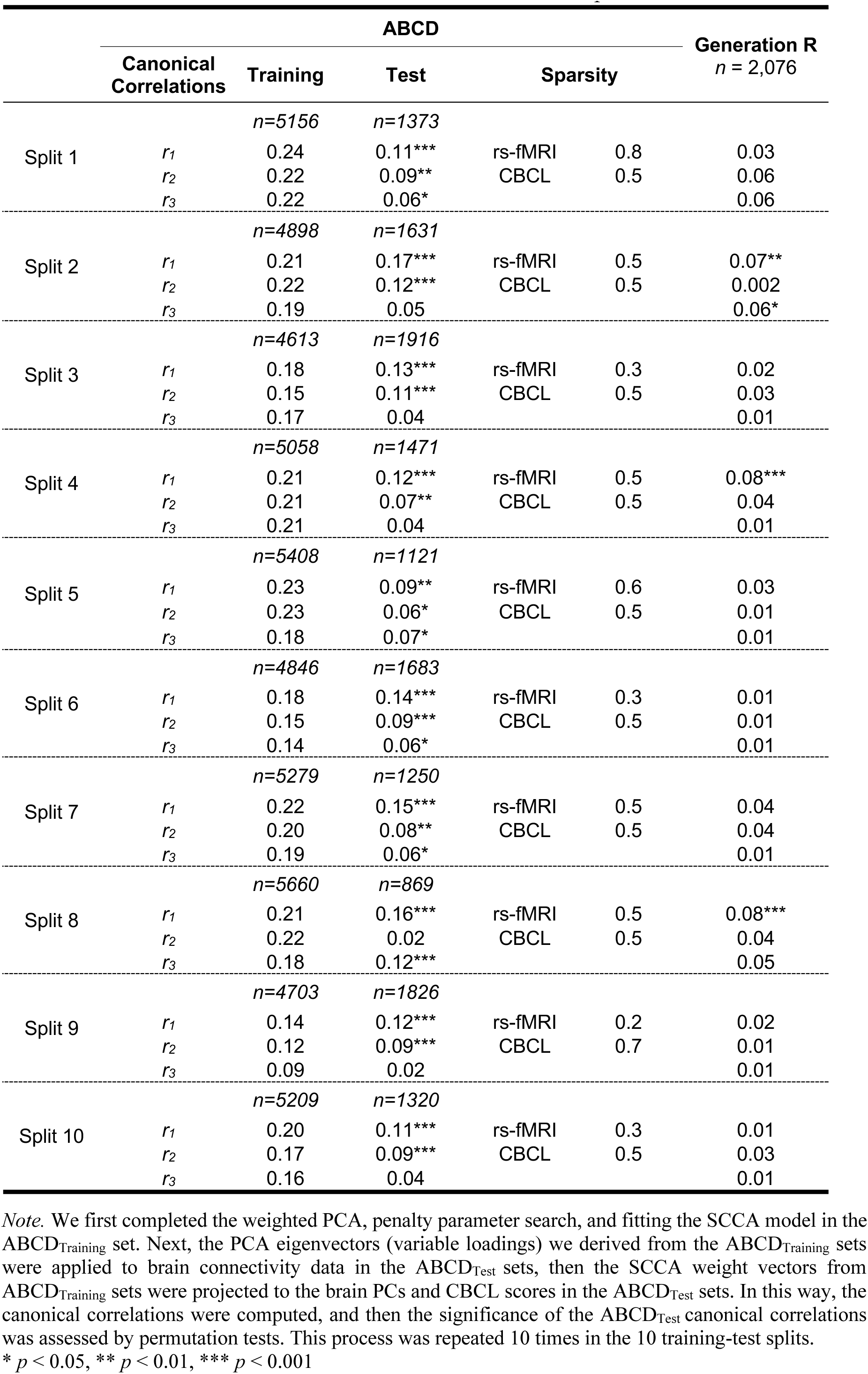
Canonical correlations in ABCD and Generation R across 10 splits.

### Out-of-sample generalizability of brain-behavior dimensions

Next, to ascertain the *out-of-sample* generalizability of the model, the remaining 3 ABCD study sites (ABCD_Test_ set, repeated 10 times) were used. By applying the eigenvectors of the weighted PCA along with the resulting weight vectors from the SCCA of the ABCD_Training_ set, the model parameters were projected onto the functional connectivity data and psychiatric symptom data from the ABCD_Test_ set. This process yielded out-of-sample canonical correlations. Overall, we found evidence that the first two canonical variates were robustly identified, and the third to a lesser extent (Table 2). The first dimension was consistently validated across the 10 splits (*r*_1_ = 0.13, *p* < .001, permutation testing; averaged across 10 splits). This brain-symptom dimension captured the correlates between attention problems and connectivity in networks involved in higher-order functions (e.g., salience and frontoparietal network)^30^, visual-spatial attention network (parietal occipital, medial parietal network)^31^, auditory, and default mode network (Figure 3a, 3d). The second dimension was evident in most of the train-test splits (*r*_2_ = 0.08, *p* < .05, permutation testing; averaged across 10 splits). This dimension delineated a relationship between aggressive/rule-breaking behaviors and connectivity patterns in similar networks involved in the first dimension, with a larger contribution from subcortical areas (e.g., hippocampus, thalamus) and ventral attention network (Figure 3b, 3e). The third dimension was observed in half of the train-test splits (*r*_3_ = 0.06, *p* < .05 permutation testing; averaged across 10 splits). Here, a correlation between withdrawn and anxious/depressed problems and connectivity patterns mostly in ventral attention, default mode networks, and subcortical areas (e.g., thalamus, amygdala) was observed (Figure 3c, 3f). Interestingly, the default mode, medial parietal, parietal occipital networks, and subcortical areas overlapped across three dimensions. Importantly, when splitting the ABCD sample into train/test sets differently (i.e., allowing all study sites to be represented in both training and testing sets), the first three canonical variates were more stable and demonstrated a smaller decrease in canonical correlations from training to test set (Supplementary Table 3). These results suggest the SCCA likely overfits when training and testing sets contain data from all ABCD study sites, and also demonstrate SCCA has the potential to ‘learn’ differences across sites (e.g., demographic differences, residual site effects).

**Figure 3.**
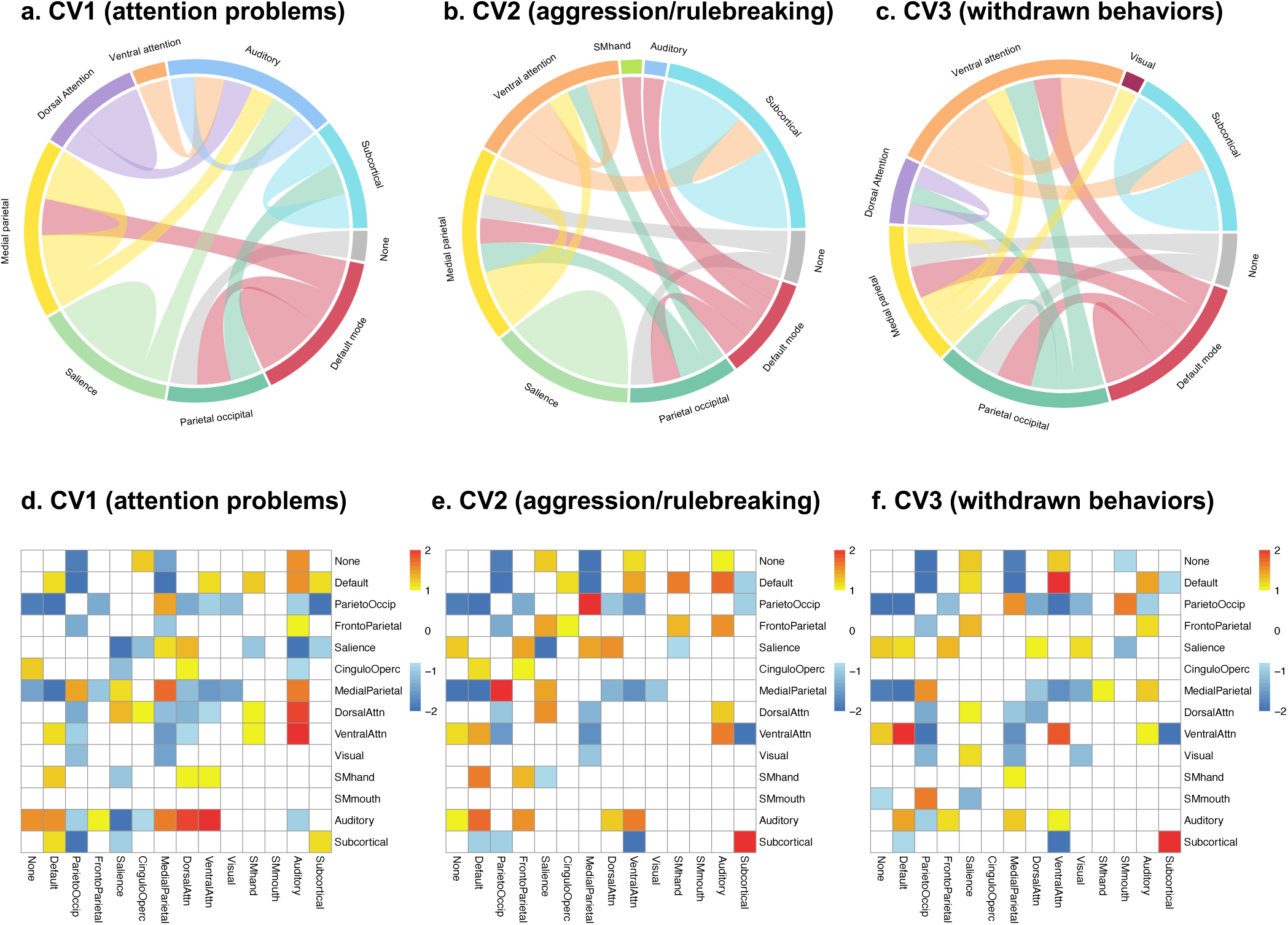
Resting-state connectivity canonical variates in ABCD. *Note.* Brain connectivity modules involved in the three identified canonical variates in ABCD. The contribution of each connectivity feature was determined by computing the correlations between the original connectivity matrix and the canonical variate scores of the brain connectivity extracted from the SCCA model (calculated by canonical loadings averaged across 10 train-test splits and the whole sample of ABCD), indicating the importance of each connectivity feature. After calculating the contribution of each connectivity feature, we summarized the contributions based on pre-assigned network modules and calculated the within and between-network loadings based on the network module analysis method in Xia, et al. (2018). **a-c.** The top 5% of the connectivity patterns that contributed most for each of canonical variate. The outer labels represent the names of network modules. The thickness of the chords showed the importance of different network modules. **d-f.** The connectivity patterns associated with the first three canonical variates. This is based on the z-scores of the within- and between-network loadings we calculated. None is the community not labeled.

### Stability of the brain-based dimensions

To further interpret the characteristics of each canonical variate and the stability of canonical loadings, 1000 bootstrap subsamples were generated in ABCD (see Methods). The variability of the first three canonical correlations, CBCL canonical loadings, and brain connectivity canonical loadings are presented in Figure 4. This again validated the three canonical variates we identified across the 10 train-test splits, showing that the loadings are relatively stable. Importantly, the three canonical correlations decreased considerably in the ABCD_Test_ set compared to the ABCD_Training_ set, especially for the second and third canonical correlations (Figure 4c). Consistent with this larger decrease of the second and third correlations, the instability of rs-fMRI canonical loadings manifested through more variability in the canonical loadings for the second and third canonical variates, while relatively stable contribution from CBCL syndrome scores was observed (Figure 4a, 4b).

**Figure 4.**
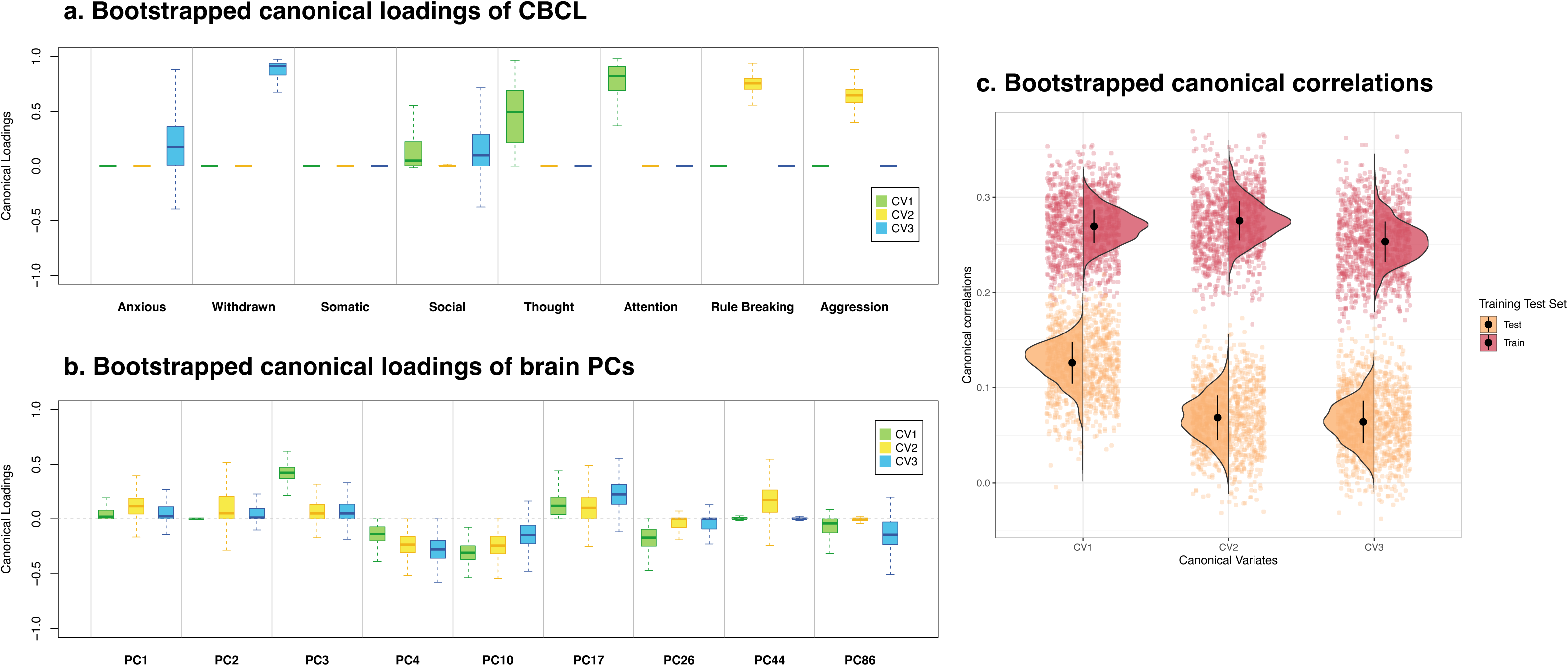
Stability and sampling variability of canonical correlations and canonical loadings in ABCD (example) *Note.* Sampling variability and important contributors for the first three canonical variates. **a.** The variability for the canonical loadings of CBCL syndrome scores across 1000 bootstrap subsamples. **b.** The variability for the canonical loadings of brain PCs across 1000 bootstrap subsamples. The PCs presented here were selected based on the intersection of top 10 most important PCs for the first three canonical variates. **c.** The variability of the first three canonical correlations in ABCD_Training_ and ABCD_Test_ set. The black dot is mean, and the vertical black line is standard deviation. Note that the bootstrap subsampling is conducted in one of the 10 train-test splits. **CV1**: canonical variate 1, **CV2**: canonical variate 2, **CV3**: canonical variate 3.

### Out-of-study generalizability in a fully independent sample

Although the ABCD Study is a multisite study, it is a highly harmonized dataset in the context of the imaging and behavioral data, and also likely has sampling characteristics that are specific and uniform across sites. Therefore, in order to test the *out-of-study* generalizability of the results we obtained in ABCD, we use the Generation R Study as an independent external validation set. The Generation R Study is a single-site population-based birth cohort in Rotterdam, the Netherlands^23^, which has ascertained a large, early adolescent sample with very similar measures as the ABCD Study. We included 2,076 children at the age of 10 with good-quality resting-state connectivity data (see Methods). We characterized two approaches of external validation. One is the commonly used ‘qualitative replication’, where the SCCA model was independently trained on Generation R and the results between cohorts were correlated. Another is the ‘gold-standard’ test, where we directly projected the SCCA model weights of the ABCD_Training_ set onto Generation R.

In the gold-standard generalizability test, the first canonical variate survived permutation tests in only 3 of the 10 train-test splits In Generation R (Table 2). No other canonical correlations survived permutation tests using the SCCA model that was trained in ABCD (*r_1_*=0.04, *r_2_*=0.03, *r_3_*=0.03, *ps* > 0.05; Supplementary Table 4). In the qualitative replication, where the SCCA was re-run in Generation R, five significant canonical variates were identified. Specifically, one canonical variate (attention problems) showed a Pearson correlation of *r*=0.79 for the CBCL canonical loadings between the two cohorts. Further, the canonical variate related to withdrawn behaviors was also similar, showing a Pearson correlation of *r*=0.67 between the CBCL canonical loadings. The canonical variate of aggressive and rule-breaking behaviors differed in the two cohorts (*r*=0.18 correlation in loadings), where it delineated aggressive and social problems in Generation R (Supplementary Figure 1). The remaining two canonical variates found in Generation R (Supplementary Figure 2a) did not overlap considerably with those observed in ABCD.

### Further exploration of brain canonical variates

One popular application of the dimensionality reduction and multimodal fusion of SCCA in neuroimaging is to utilize the brain canonical variates as the input for other statistic models or clustering algorithms^4, 17^. Thus, in a final step, we first explored whether the identified brain canonical variates were associated with cognitive ability in ABCD (see Methods). We found that the first and second brain canonical variates (associated with attention problems and aggressive/rule-breaking behaviors, respectively) were related to fluid and crystallized intelligence, matrix reasoning scores, and total cognition scores, while the third brain canonical variate (withdrawn behaviors and anxious/depression) was only marginally associated with matrix reasoning scores (Supplementary Table 5).

## Discussion

Several studies have highlighted the intriguing potential of multivariate brain-behavior associations, but the lack of replicability of results has hampered the identification of robust neurobiological mechanisms underlying psychiatric problems^3, 17^. To maximize the robustness and generalizability of brain-behavior associations in a fully independent sample, which is largely absent in previous research in the psychiatric neuroimaging literature, the present study moved beyond solely testing *out-of-sample* generalizability in a single cohort, to evaluating *out-of-study* generalizability in a fully external cohort. Robust multivariate brain-psychiatric symptom associations in children were observed, however, the gold-standard test of generalizability in an external cohort was largely negative. While these results reinforce previous work demonstrating the potential for brain-based dimensions of psychiatric problems, they also highlight the deep-rooted problem of poor generalizability in psychiatric neuroimaging studies.

In ABCD, we identified two brain-symptom dimensions that were consistently validated in the out-of-sample test sets, indicating robust *within-study (internally valid)* multivariate brain-symptom associations. The first two brain-symptom dimensions center on externalizing problems (attention problems, aggressive and rule-breaking behaviors). Several connectivity networks loading highly on these dimensions, such as salience, default mode, parietal occipital, and medial parietal networks, have been shown to be involved in attention deficit hyperactivity disorder (ADHD)^32, 33^. These networks have also been implicated in deficits of top-down executive control, attention, and spatial working memory in children with ADHD^31–34^. The third brain-symptom dimension characterizes internalizing problems, representing behaviors such as withdrawal, anxiety, and depression^35^. Consistent with previous findings in adults and children^36–38^, the default mode and ventral attention networks manifested as the major contributors, which are related to emotional dysregulation and impaired reorientation of attention^36, 39^. The three identified brain-based dimensions were further validated by their associations with child cognitive ability, which is in line with results in behavioral studies showing associations between externalizing/internalizing problems and intelligence^40^.

While we discovered three brain-symptom dimensions in ABCD, the out-of-study generalizability in Generation R presented a complex picture. Psychiatric neuroimaging studies employ varying approaches to test generalizability, and thus demonstrate varying degrees of external validity. One commonly used approach consists of repeating the analyses in data that were previously ‘unseen’, and then qualitatively (and to some extent, quantitatively) comparing results. In the present study, we observed similar behavioral dimensions when training the SCCA model independently in Generation R. Two dimensions were highly robust in ABCD and observed in the qualitative external replication, lending support for reasonable internal validity of these brain-behavior dimensions. Therefore, the results are convincing in the general context of underlying dimensional neurobiology. However, even though this route of ‘replication’ is a valuable way to demonstrate whether the brain-behavior associations *exist* from an empirical perspective, precisely how one can define a ‘successful’ replication based on the qualitative or quantitative *similarities* between results remains a non-trivial challenge for the field.

Importantly, the more robust, gold-standard generalizability test yielded less optimistic results. In clinical prediction, which is arguably the primary goal of machine-learning models in psychiatric neuroimaging, a ‘gold-standard’ test demonstrates a much higher degree of real-world generalizability. The lack of this degree of generalizability in an external, independent sample suggests that the dimensions cannot be applied to other datasets as a potential biomarker. In the subsequent paragraphs, we will delve into the potential explanations of the challenges in this generalizability test, and then ultimately provide recommendations on how to improve out-of-study generalizability.

First, the multivariate method we utilized, CCA, is highly prone to overfitting and instability^18, 41^ and requires a large sample size to obtain sufficient statistical power^20^. In our study, the sample size of Generation R (*n*=2,076) might not be large enough to capture the true associations. However, Generation R is similar in size to our ABCD_Test_ set (*n* ∼ 1500), where we successfully validated associations. Second, the vast majority of previous studies drew from clinical samples with a specific diagnosis, such as depression, psychosis, and ADHD^2, 17, 42^. Focusing on the general population, rather than clinical samples, might dilute associations. However, the utility of dimensional assessments of symptoms is well-known and has several advantages to problems in clinical, case-control designs. Third, rs-fMRI data has intrinsically high inter-individual variability than other brain measures in psychiatry^43^, thus extracting clinically important signals on an individual basis is difficult and generalizability across cohorts could be especially challenging.

Another important reason that we consider is that the rs-fMRI data in ABCD and Generation R could not be fully harmonized. Nevertheless, our results show that this is unlikely to be the main driver of low generalizability, and rather that there is a strong *site effect*. Using the gold-standard generalizability test, we observed several generalizable canonical correlations in Generation R despite the two cohorts being independent in many aspects. Moreover, even within ABCD, a fully harmonized cohort in terms of imaging acquisition and image preprocessing, there was a significant drop in canonical correlations from the training to test sets of 50% or more. Importantly, when the train-test split disregards the site information (e.g., random), we observed less degradation of performance. Taken together, the low generalizability of our models is likely driven by factors inherently embedded in different study sites that cannot be completely accounted for by data harmonization. Model failure is thus intertwined with the difference of other confounding factors which are distinct across cohorts^7, 44^.

A few limitations of the study should be noted. First, we only applied SCCA in our analysis. Other multivariate methods were not examined. Yet, CCA is one of the most widely used techniques, and other multivariate methods have been found to be sensitive to similar problems of generalizability^3, 5^. Second, ABCD and Generation R were not fully harmonized in terms of imaging acquisition and processing. However, as discussed above, a clear site effect was observed even within ABCD where the data were fully harmonized, and thus it is unlikely that the harmonization will lead to considerable differences. As there will never be a situation where data across the world can be perfectly harmonized, it is crucial that we identify methods that are less sensitive to differences across studies.

## Conclusions

In summary, the utilization of SCCA enabled us to discover robust brain-symptom associations. The results offer substantial room for optimism about using multivariate methods in brain-behavior association studies. Future studies could further explore whether these brain-based dimensions could inform more targeted prevention, detection, and intervention of child psychiatric disorders. However, to achieve this goal of clinical utility, future studies must test results in fully external validation sets. Further, more robust, gold-standard generalizability tests are crucial for the clinical translation of results (e.g., applying model coefficients from one study directly to an external validation set). Finally, in addition to data harmonization, hidden confounders across sites or studies should be considered. Recent advances in methods of accommodating site variations might also considerably boost generalizability and reduce the site differences^45^.

## Methods

### Study population

This study is embedded in two prospective cohorts of child development, the ABCD study^24^ and the Generation R Study^23^.

#### The ABCD Study

The ABCD study assesses brain development from pre-adolescence to adulthood and was conducted across 21 study sites in the United States. Children aged 9-10 years were recruited as baseline and the sample is epidemiologically-informed^24^. In the ABCD cohort, resting-state functional magnetic resonance imaging (rs-fMRI) was obtained through the ABCD-BIDS Community Collection (ABCC), a community-shared ABCD neuroimaging dataset that is continually updated (https://collection3165.readthedocs.io). Both the rs-fMRI data and the behavioral assessments (data release 4.0) were retrieved from the baseline visit data of children aged 9-11 years old. Details of the study design and exclusion criteria are described in previous reports^24^. Of the 9,400 children whose rs-fMRI data were available, we excluded 1,398 children who failed the quality control of the resting-state connectivity data (see below), 303 children with incidental findings, and 23 children with any missingness in behavioral measures and covariates. For families with multiple participants, one twin or sibling was randomly included (1,147 excluded). Accordingly, data from 6,529 participants were available for analysis in ABCD.

#### Generation R

The Generation R Study is a population-based birth cohort in Rotterdam, the Netherlands. Rs-fMRI data and behavioral assessments were obtained as part of the age-10 data collection which began in 2013^23^. Among the 3,992 children who were scanned with MRI, 3,170 completed rs-fMRI scanning. We excluded children as a result of the image quality assurance protocol (see below, *n*=583), and children with higher than 25% missing values in the behavioral assessments (*n*=446). After randomly including one twin or sibling (*n*=65), 2,076 participants were included in the final sample for analysis.

### Measures

#### Child psychiatric symptoms

Child psychiatric symptoms were assessed using the Child Behavioral Checklist (school-age version)^46^. The CBCL is a 118-item caregiver report with eight syndrome scales (anxious/depressed, withdrawn/depressed, somatic, social, aggressive, rule-breaking, thought, and attention problems), assessing child internalizing and externalizing problems. Internalizing problems reflect a variety of inner-directed symptoms, such as anxiety, withdrawal, or depression, while externalizing problems incorporate outer-directed symptoms, such as aggression and rule-breaking behaviors^47^. The CBCL was administered in both cohorts and the primary caregivers answered 118 items on a three-point scale (not true, sometimes true, very often or always true) for problems in the past six months. Raw sum scores of the syndrome scales were utilized in the current study, with higher scores representing more problems.

#### fMRI pre-processing

The ABCD data sets were retrieved from ABCC. In ABCC, the BIDS data were preprocessed with the abcd-hcp-pipeline (https://github.com/DCAN-Labs/abcd-hcp-pipeline), a modification and extension of the Human Connectome Project (HCP) Minimal Preprocessing Pipelines^48, 49^. Structural data undergo a multi-step pre-processing procedure first, including brain extraction, denoising, and B_1_-inhomogeneity (bias field) correction (“pre-FreeSurfer” phase). Next, structural scans are processed through the FreeSurfer software suite (“FreeSurfer” phase). Nonlinear registration using the ANTs toolbox is then applied to warp structural data to MNI space (6^th^ Generation MNI ICBM 152 supplied with FSL^50^, “post-FreeSurfer” phase). Resting-state data were then intensity normalized, corrected for geometric distortions, undergo volume realignment to correct and assess head motion, and aligned first to the structural scan and then to the MNI template by concatenating with the previously determined warp (“Vol” phase). Lastly, data were projected to surface-based space (32k fs_LR).

In the Generation R study, rs-fMRI data were preprocessed using the FMRIPrep pipeline (version 20.1.1 singularity container)^51^. Briefly, structural MRI data first underwent intensity normalization to account for B_1_-inhomogeneity and brain extraction, followed by nonlinear registration to MNI space and FreeSurfer processing. Functional MRI data first underwent volume realignment with MCFLIRT (FSL). BOLD runs were then slice-time corrected with 3dTshift (AFNI), followed by co-registration to the corresponding T1w reference. Spatial normalization to the ICBM 152 Nonlinear Asymmetrical template version 2009c^52^ was conducted through nonlinear registration with the antsRegistration tool of ANTs v2.1.0^50^, using the above-mentioned T1w reference in the registration scheme. Data were ultimately resampled to Cifti format in 32k fs_LR surface space.

#### Parcellation and whole-brain connectivity estimation

Within ABCD, the resting-state functional connectivity matrices were processed using the DCANBOLDProcessing (DBP) resting-state fMRI processing tools (https://github.com/DCAN-Labs/dcan_bold_processing). This consisted of applying a respiratory filter, flagging volumes with FD > 0.3mm as contaminated with motion, demeaning and detrending of data, and denoising of data by regressing out whole brain, ventricular and white matter (and their derivatives) signals, and finally bandpass filtered between 0.008 and 0.1 Hz to avoid potential aliasing of the time series signal. The processed functional data was used to generate correlation matrices using Pearson correlation, followed by Fisher z-transformation (https://collection3165.readthedocs.io/en/stable/pipeline/). Following the instruction of the ABCC collection, we downloaded the available functional connectivity matrices that were calculated and labeled using the Gordon cortical parcels^29^ and FreeSurfer subcortical segmentation^53^. This yielded 352 distinct parcels consisting of 333 cortical and 19 subcortical regions.

Within Generation R, whole-brain functional connectivity matrices were calculated and mapped onto the same 333 cortical and 19 subcortical regions with ABCD. Similar to ABCD, the extracted time series were adjusted for CSF and white matter signals (not global signal), low-frequency temporal regressors for high pass temporal filtering, and the ICA AROMA components related to motion artifacts. Next, we removed the first 4 volumes of each subject to ensure magnetic stabilization, then BOLD signals were averaged across all voxels in each cortical and subcortical region. Connectivity estimation was the same across cohorts, including the Pearson correlation that was applied to estimate the temporal dependence between the residualized regional time series and Fisher z-transformation, resulting in a symmetric 352 × 352 functional connectivity matrix for each participant.

#### Quality controls of the scans

In the ABCC data sets, only data that passed the initial acquisition Data Analysis Imaging Center (DAIC) quality control were included. At the time of scanning, quality control was performed by scan operators with a binary pass or fail. Images were also visually inspected for motion and other major artifacts. Automated measures of quality control (e.g., FD and also temporal SNR) were also applied. Participants were excluded based on the recommended guidelines (imgincl_rsfmri_include = 1), which involve raw and postprocessing quality control, passed FreeSurfer QC, had more than 375 rs-fMRI frames after censoring, and other cut-off scores (see ABCD Release 4.0), for a total of 1,398 participants excluded due to poor quality. In addition, we excluded 303 participants with clinically relevant incidental findings.

In Generation R, the following exclusion criteria were applied to screen eligible participants (1) scans with major artifacts (e.g., dental retainers, or other scan-related artifacts) (2) scans lacking whole-brain coverage (e.g., missing large portions of the cerebrum or cerebellum from the field of view) (3) scans with excessive motion (mean framewise displacement (FD) higher than 0.25 mm or having more than 20% of the volumes with an FD higher than 0.2 mm)^54^. Moreover, the accuracy of co-registration was visually inspected by merging all co-registered images into a single 4D Nifti image and scrolling through the images. 583 scans with poor quality were excluded in total.

#### Covariates

In ABCD, child age, sex, race/ethnicity, parental education, and data collection site were used as covariates. Demographic information (child age, sex, race/ethnicity, and parental education) were assessed by parent-report questionnaires. The original 21-category parental education was recoded into three categories to make it comparable with Generation R: 1^st^ to 12^th^ grade, high school/GED/college, and Bachelor’s degree or higher.

In Generation R, similar covariates were included except for study sites, including age of children when undergoing the MRI scanning, sex, child national origin, and maternal education. Child national origin was defined based on the birth country of the parents and was coded into three categories: Dutch, non-Dutch European, and non-European^55^. Maternal education, an indicator of socioeconomic status, was recoded into three categories: maximum of three years secondary school, more than three years general secondary school; intermediate vocational training, and Bachelor’s degree or higher^56^. Missing values were imputed by using Expectation-Maximization imputation as the proportion of missing values was smaller than 1% of the current Generation R data set^57^.

#### Child cognitive ability

Child cognitive ability data was retrieved from NIH Toolbox age-corrected standard scores of fluid intelligence (adaptive problem-solving), crystallized intelligence (knowledge acquisition from experience), total cognition scores (overall cognition composite scores), and matrix reasoning scaled scores (non-verbal reasoning) from the Wechsler Intelligence Scale for Children-V (data release 4.0)^58, 59^.

### Statistical analysis

#### Analysis framework

The current study implemented a multiple hold-out framework that aims to increase the generalizability of the analysis^28^. We used ABCD as the discovery set (*n*=6,529), in which all analyses were conducted (trained) and tested. The ABCD discovery set was randomly split into a training set consisting of 18 sites (ABCD_Training_) and a test set consisting of 3 sites (ABCD_Test_). In this way, subjects in the ABCD_Training_ and ABCD_Test_ sets were entirely from different sites, approaching the true out-of-sample context (Figure 1). To reduce sampling biases, the split procedure was repeated 10 times, resulting in 10 pairs of independent train-test sets. Importantly, the analyses in ABCD_Training_ sets and ABCD_Test_ sets were fully separated to safeguard the results from data leakage (Figure 1). Specifically, the model was trained in ABCD_Training_ set, where the dimensionality reduction (see ***weighted PCA below***) was done, and the performance of the hyperparameter of the models were selected in 100 further random splits of training (80% of ABCD_Training_ set) and validation set (20% of ABCD_Training_ set). After fitting the model with the optimal hyperparameters in the ABCD_Training_ set, *out-of-sample* model generalizability was evaluated in the ABCD_Test_ set. In a final step, Generation R, which has ascertained a large early adolescent sample with very similar measures, was used as an independent external validation set (*n*=2,076). We characterized two approaches of external validation (see ***Out-of-study generalizability test in Generation R***), allowing us to estimate the *out-of-study* generalizability of the findings from ABCD. Moreover, we did several explorations of the identified brain canonical variates in ABCD. First, we tested whether the identified brain canonical variates were associated with child cognitive ability at the age of 10. Second, we investigated whether we could find distinct subgroups/clusters of children based on the identified brain canonical variates.

#### Dimensionality reduction

Prior to SCCA analysis, the upper triangle of the 352 × 352 functional connectivity matrix was flattened, resulting in 61,776 connectivity features for each participant. Connectivity values were residualized to ensure the above-mentioned covariates did not influence the results^16^. As the high-dimensional nature of the connectivity features could lead to considerable overfitting in SCCA, weighted principal component analysis (PCA) was applied to reduce the connectivity features into principal components (PCs) that aggregated the information of the data^60^. This PCA-CCA framework has been used extensively and has shown good performance^15^.

While traditional PCA only considers the structure of the brain data, the weighted PCA uses the relationship between the brain and behavioral data in dimensionality reduction to identify a relatively small number of PCs carrying information from the phenotypes of interest^60^. This ensures the variability in the functional connectivity data most related to behavioral and emotional problems will be captured in the PCs. To achieve this, we rescaled the connectivity data according to a rank-based weighting scheme, which depends on the sum of CBCL scores. The weight assigned to each subject was determined by the rank of their total CBCL score. The rank-based pre-weights were calculated as follows:

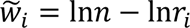

Where *n* is the number of data points and *r* is the ranking. We normalized the pre-weights by 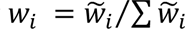, and the original connectivity data was demeaned and adjusted with the corresponding normalized weights. We then submitted the adjusted connectivity matrix to PCA, and the eigenvectors (variable loadings) of PCA were extracted and multiplied with the original connectivity matrix, resulting in a new, dimensionally reduced weighted connectivity matrix. To further protect against overfitting in subsequent analyses, a selection of PCs was made, namely the first 100 principal components^15^.

#### Sparse Canonical Correlation Analysis

##### Sparse CCA

To delineate multivariate relationships between functional connectivity and child psychiatric problems, we applied sparse CCA (SCCA), an unsupervised learning technique that can simultaneously evaluate the relationships between two sets of variables from different modalities^4^. SCCA imposes both l_1_-norm and l_2_-norm penalty terms, an elastic net regularization combining the LASSO and ridge penalties, to high-dimensional data sets and achieves sparsity of the solution^61^. This method is more stable and does not have the main constraint of classic CCA: the number of observations should be larger than the number of variables^16^. Specifically, given two matrices, Χ_𝑛×𝑝_ and Y_𝑛×𝑞_, where 𝑛 is the number of participants, 𝑝 and 𝑞 are the number of variables (e.g., CBCL scores and brain PCs, respectively), SCCA aims to find 𝑢 and 𝑣 (canonical loading matrices) that maximize the covariance between 𝑋𝑢 and 𝑌𝑣. 𝑋𝑢 and 𝑌𝑣 are canonical variates that are the low dimensional representation of brain and behavioral measures.

##### Selection of penalty parameters

Using the extracted 100 brain PCs after dimensionality reduction, we first determined the optimal penalty parameters before fitting the SCCA. In order to identify the best set of penalty parameters for the SCCA of functional connectivity and behavioral features, we used a repeated resampling procedure of the ABCD_Training_ set^27, 28^ (Figure 1). Specifically, we first split the ABCD_Training_ set further into penalty parameter training (80%) and validation set (20%) 100 times, resulting in 100 pairs of training and validation sets. Next, a grid search between 0 and 1 with increments of 0.1 was used to determine the combination of penalty parameters (l_1_ and l_2_) that show the best performance^16^. For each combination of penalty parameters, we fitted the SCCA model in the training set, projected the canonical loadings extracted from the training set (𝑢 and 𝑣) on the validation set, and then calculated the canonical correlations. The optimal combination of penalty parameters was chosen based on the highest first canonical correlation of the validation set averaged across 100 splits^61^.

##### Fitting SCCA model and significance test

After the selection of optimal penalty parameters, the SCCA model was fitted to ABCD_Training_ set with the chosen parameters. The resulting weight vectors (canonical loadings) from ABCD_Training_ set were then projected onto brain PCs and CBCL scores of ABCD_Test_ set (after first deriving brain PCs in the ABCD_Test_ set by applying the eigenvectors of the weighted PCA from ABCD_Training_ set). This process yielded the canonical correlations in the ABCD_Test_ set, reflecting the within-cohort out-of-sample generalizability of the SCCA model. To determine the statistical significance of each canonical correlation, a permutation testing procedure was applied both in the ABCD_Training_ and ABCD_Test_ sets. In the permutation test, the rows of the behavioral data were shuffled to disrupt the relationship between the brain connectivity features and the behavioral features, while the brain connectivity matrix was held constant^18^. We performed 2,000 permutations, building a null distribution of each canonical correlation. The *p*-value of the permutation test is defined as the number of null correlations that exceeded the correlations estimated on the original, un-shuffled dataset. The same set of penalty parameters was used in each permutation. Only canonical variates surviving permutation testing (*p* < 0.05) were selected for further analysis.

##### Stability of SCCA model

The classical CCA has been found to be unstable at times and fails to converge when the samples-to-feature ratio is small^20^. To investigate the sampling variability of the canonical loadings and inspect the features that consistently contributed to each canonical variate in the SCCA model, 1,000 bootstrapping subsamples (sample with replacement) were generated. The distribution of canonical loadings in this procedure allows us to inspect the stability and sampling variability of the SCCA model. This was done in one randomly selected train-test split. As arbitrary axis rotation could be induced by bootstrapping, leading to the changes of the order of canonical variates and sign of the canonical weights, we matched the order of canonical variates based on the CBCL loadings we derived from the original datasets^16^.

#### Associations with cognitive ability

To further validate the canonical variates we found, we tested whether the identified brain canonical variates were associated with child cognitive ability at the age of 10 in the ABCD cohort. We separately modeled the relationship between each significant canonical variate of brain connectivity and the cognitive ability of the participants with linear regression models adjusted for all covariates.

#### Out-of-study generalizability in Generation R

CCA is vulnerable to overfitting and the generalizability of the canonical variates should be carefully investigated^20, 28^. In the current study, we tested the generalizability of the findings from the ABCD discovery set in an external validation set: Generation R. We utilized two approaches to test the generalizability: the qualitative replication and the gold-standard test. In the qualitative replication, a common practice in current psychiatric neuroimaging studies, the SCCA model was independently trained on Generation R, yielding another set of canonical loadings. The Pearson correlation between the two sets of canonical loadings (ABCD and Generation R) was calculated as a quantitative indicator of generalizability. Similar to what is described above for ABCD, in another, more standard practice in machine learning studies, the ‘gold-standard’ test, we projected the SCCA canonical loadings of ABCD_Training_ set directly on Generation R. The canonical correlations were ultimately calculated and assessed with permutation testing.

#### Data availability

The ABCD data reported in this paper are openly available upon approval from the NDA Data Access Committee. The ABCD data came from ABCD collection 3165 (ABCD-BIDS Community Collection (ABCC), https://collection3165.readthedocs.io) and the Annual Release 4.0 (https://doi.org/10.15154/1523041).

The Generation R datasets generated and/or analyzed during the current study may be made available upon request to the Director of the Generation R Study, Vincent Jaddoe, in accordance with the local, national, and European Union regulations.

#### Code availability

All analysis code is publicly available in the following GitHub repository: (https://github.com/EstellaHsu/Brain_dimensions_ABCD_GenR).

## Data Availability

The ABCD data reported in this paper are openly available upon approval from the NDA Data Access Committee. The ABCD data came from ABCD collection 3165 (ABCD-BIDS Community Collection (ABCC), https://collection3165.readthedocs.io) and the Annual Release 4.0 (https://doi.org/ 10.15154/1523041).
The Generation R datasets generated and/or analyzed during the current study may be made available upon request to the Director of the Generation R Study, Vincent Jaddoe, in accordance with the local, national, and European Union regulations.

## Acknowledgements

Data used in the preparation of this article were obtained from the Adolescent Brain Cognitive Development^SM^ (ABCD) Study (https://abcdstudy.org), held in the NIMH Data Archive (NDA). This is a multisite, longitudinal study designed to recruit more than 10,000 children age 9-10 and follow them over 10 years into early adulthood. The ABCD Study® is supported by the National Institutes of Health and additional federal partners under award numbers U01DA041048, U01DA050989, U01DA051016, U01DA041022, U01DA051018, U01DA051037, U01DA050987, U01DA041174, U01DA041106, U01DA041117, U01DA041028, U01DA041134, U01DA050988, U01DA051039, U01DA041156, U01DA041025, U01DA041120, U01DA051038, U01DA041148, U01DA041093, U01DA041089, U24DA041123, U24DA041147. A full list of supporters is available at https://abcdstudy.org/federal-partners.html. A listing of participating sites and a complete listing of the study investigators can be found at https://abcdstudy.org/consortium_members/. ABCD consortium investigators designed and implemented the study and/or provided data but did not necessarily participate in the analysis or writing of this report. This manuscript reflects the views of the authors and may not reflect the opinions or views of the NIH or ABCD consortium investigators. The Generation R Study is supported by Erasmus MC, Erasmus University Rotterdam, the Rotterdam Homecare Foundation, the Municipal Health Service Rotterdam area, the Stichting Trombosedienst & Artsenlaboratorium Rijnmond, the Netherlands Organization for Health Research and Development (ZonMw), and the Ministry of Health, Welfare and Sport. Neuroimaging data acquisition was funded by the European Community’s 7th Framework Program (FP7/2008-2013, 212652, Nutrimenthe).

Netherlands Organization for Scientific Research (Exacte Wetenschappen) and SURFsara (Snellius Compute Cluster, www.surfsara.nl) supported the Supercomputing resources. Authors are supported by an NWO-VICI grant (NWO-ZonMW: 016.VICI.170.200 to HT) for HT, BX, and the Sophia Foundation S18-20, and Erasmus MC Fellowship for RLM. We gratefully acknowledge the participants, general practitioners, hospitals, midwives, and pharmacies in Rotterdam who contributed to the study.

## Author contributions

Based on the CRediT role taxonomy (https://credit.niso.org/): conceptualization (HT, RLM, JF, BX), data curation (RLM, BX, ML, PC (ABCD)), formal analysis (BX), funding acquisition (HT, RLM), investigation (BX), methodology (BX, HT, RLM, HW, GB), project administration (RLM, HT, BX), resources (RLM), software (BX, LDA), supervision (RLM, HT), validation (BX), visualization (BX), writing – original draft (BX), writing – review and editing (BX, RLM, HT, LDA, HW, AM, PC, JF, BTC, MB, ML, GB).

