## Supplementary Information for "Multivariate brain-based dimensions of child psychiatric problems: degrees of generalizability"

1. Supplementary tables 1-5
2. Supplementary figures 1-2

**Supplementary Table 1**

*MRI acquisition parameters in ABCD*

| | Matrix | Slices | FOV | TR (ms) | TE (ms) | TI (ms) | Flip Angle (deg) | MultiBand Acceleration | Acquisition Time |
|---|---|---|---|---|---|---|---|---|---|
| **Siemens** | | | | | | | | | |
| **T1** | 256 × 256 | 176 | 256 × 256 | 2500 | 2.88 | 1060 | 8 | Off | 7:12 |
| **fMRI** | 90 × 90 | 60 | 216 × 216 | 800 | 30 | N/A | 52 | 6 | |
| **Philips** | | | | | | | | | |
| **T1** | 256 × 256 | 225 | 256 × 240 | 6.31 | 2.9 | 1060 | 8 | Off | 5:38 |
| **fMRI** | 90 × 90 | 60 | 216 × 216 | 800 | 30 | N/A | 52 | 6 | |
| **GE** | | | | | | | | | |
| **T1** | 256 × 256 | 208 | 256 × 256 | 2500 | 2 | 1060 | 8 | Off | 6:09 |
| **fMRI** | 90 × 90 | 60 | 216 × 216 | 800 | 30 | N/A | 52 | 6 | |

*Note.* Parameters retrieved from the ABCC collection (https://collection3165.readthedocs.io/en/stable/inputs/).

**Supplementary Table 2**

*MRI acquisition parameters in Generation R*

| | Matrix | Slices thickness (mm)/ Number of slices | FOV | TR (ms) | TE (ms) | TI (ms) | Flip Angle (deg) |
|---|---|---|---|---|---|---|---|
| **GE 750** | | | | | | | |
| **T1** | 220 × 220 | 1.0/230 | 220 × 220 | 8.77 | 3.4 | 600 | 10 |
| **fMRI** | 64 × 64 | 4.0/36 | 216 × 216 | 1760 | 30 | N/A | 85 |

*Note.* T1-weighted images were obtained using a coronal inversion recovery fast spoiled gradient recalled sequence using ARC acceleration (GE option BRAVO). Rs-fMRI data were obtained using an interleaved axial echo planar imaging sequence. Total duration of the resting-state scan was 5 minutes 52 seconds for each child.

**Supplementary Table 3**

*Canonical correlations in training and test sets of ABCD across 10 splits*

*(Train-test split with pooled multisite data)*

| | Canonical Correlations | Training | Test | Sparsity | |
|---|---|---|---|---|---|
| Split 1 | $r_1$ | 0.20 | 0.10*** | Rs-fMRI | 0.4 |
| | $r_2$ | 0.18 | 0.06* | CBCL | 0.5 |
| | $r_3$ | 0.19 | 0.10*** | | |
| Split 2 | $r_1$ | 0.18 | 0.13** | Rs-fMRI | 0.4 |
| | $r_2$ | 0.17 | 0.09* | CBCL | 0.5 |
| | $r_3$ | 0.15 | 0.11** | | |
| Split 3 | $r_1$ | 0.18 | 0.12*** | Rs-fMRI | 0.4 |
| | $r_2$ | 0.17 | 0.06* | CBCL | 0.5 |
| | $r_3$ | 0.17 | 0.07* | | |
| Split 4 | $r_1$ | 0.16 | 0.12*** | Rs-fMRI | 0.3 |
| | $r_2$ | 0.16 | 0.09** | CBCL | 0.5 |
| | $r_3$ | 0.14 | 0.10** | | |
| Split 5 | $r_1$ | 0.19 | 0.16*** | Rs-fMRI | 0.5 |
| | $r_2$ | 0.19 | 0.06* | CBCL | 0.5 |
| | $r_3$ | 0.16 | 0.11*** | | |
| Split 6 | $r_1$ | 0.15 | 0.15*** | Rs-fMRI | 0.3 |
| | $r_2$ | 0.14 | 0.07* | CBCL | 0.6 |
| | $r_3$ | 0.09 | 0.07** | | |
| Split 7 | $r_1$ | 0.17 | 0.12*** | Rs-fMRI | 0.4 |
| | $r_2$ | 0.14 | 0.06* | CBCL | 0.5 |
| | $r_3$ | 0.15 | 0.10*** | | |
| Split 8 | $r_1$ | 0.17 | 0.13*** | Rs-fMRI | 0.3 |
| | $r_2$ | 0.14 | 0.06* | CBCL | 0.5 |
| | $r_3$ | 0.15 | 0.08** | | |
| Split 9 | $r_1$ | 0.18 | 0.10*** | Rs-fMRI | 0.4 |
| | $r_2$ | 0.16 | 0.06* | CBCL | 0.5 |
| | $r_3$ | 0.16 | 0.07** | | |
| Split 10 | $r_1$ | 0.18 | 0.13*** | Rs-fMRI | 0.3 |
| | $r_2$ | 0.15 | 0.07** | CBCL | 0.5 |
| | $r_3$ | 0.15 | 0.09** | | |

*Note.* We pooled the data from 21 sites together and randomly split it into a training set ($ABCD_{Training}$, 80% of the data) and a test set ($ABCD_{Test}$, 20% of the data). The canonical correlations in the $ABCD_{Test}$ sets were calculated by applying the weight vectors obtained from the $ABCD_{Training}$ set SCCA model to the test sets. This process was repeated 10 times in the 10 training-test splits.
* $p < 0.05$, ** $p < 0.01$, *** $p < 0.001$

### Supplementary Table 4

*Failed gold-standard generalizability test in Generation R*

| Canonical correlations | ABCD | | Generation R |
|---|---|---|---|
| | training set | test set | |
| $r_1$ | 0.20 | 0.13*** | 0.04 |
| $r_2$ | 0.19 | 0.08** | 0.03 |
| $r_3$ | 0.17 | 0.06* | 0.03 |

*Note.* Canonical correlations in ABCD were averaged across the 10 train-test splits. $r_1$ : *** $p < 0.001$ across all 10 train-test splits. $r_2$: ** $p < 0.01$ in 8 train-test splits. $r_3$: * $p < 0.05$ in 5 train-test splits. $r_{1\ Generation\ R}$: p < 0.01 in 3 train-test splits.

**Supplementary Table 5**

*Correlations between the first three brain canonical variates and cognitive ability (n=5,269)*

| | **Fluid intelligence** | | **crystallized intelligence** | | **matrix reasoning** | | **total cognition** | |
|---|---|---|---|---|---|---|---|---|
| | B (95% CI) | *P* | B (95% CI) | *p* | B (95% CI) | *p* | B (95% CI) | *p* |
| **CV1** | 0.05 [0.02, 0.08] | < .001 | 0.06 [0.02, 0.07] | < .001 | 0.03 [0.01, 0.06] | 0.01 | 0.06 [0.03, 0.08] | <.001 |
| **CV2** | 0.03 [0.01, 0.06] | 0.01 | 0.04 [0.02, 0.07] | < .001 | 0.04 [0.01, 0.06] | 0.003 | 0.05 [0.02, 0.07] | <.001 |
| **CV3** | 0.02 [-0.01, 0.04] | 0.22 | 0.02 [-0.01, 0.04] | 0.22 | 0.03 [0.003, 0.05] | 0.03 | 0.02 [-0.004, 0.04] | 0.11 |

*Note*. Separate linear regression analysis of cognitive ability and the first three brain canonical variates. Betas are standardized. All the models were adjusted for child age, child sex, race/ethnicity, parental education, and scanning sites. After excluding the participants with missing values in any of the four cognitive abilities, the final sample size of this analysis is 5,269. **CV1**: brain canonical variate 1, **CV2**: brain canonical variate 2, **CV3**: brain canonical variate 3.

### Supplementary Figure 1

*CBCL canonical loadings in ABCD and Generation R in qualitative replication*

**a. ABCD** **b. Generation R**

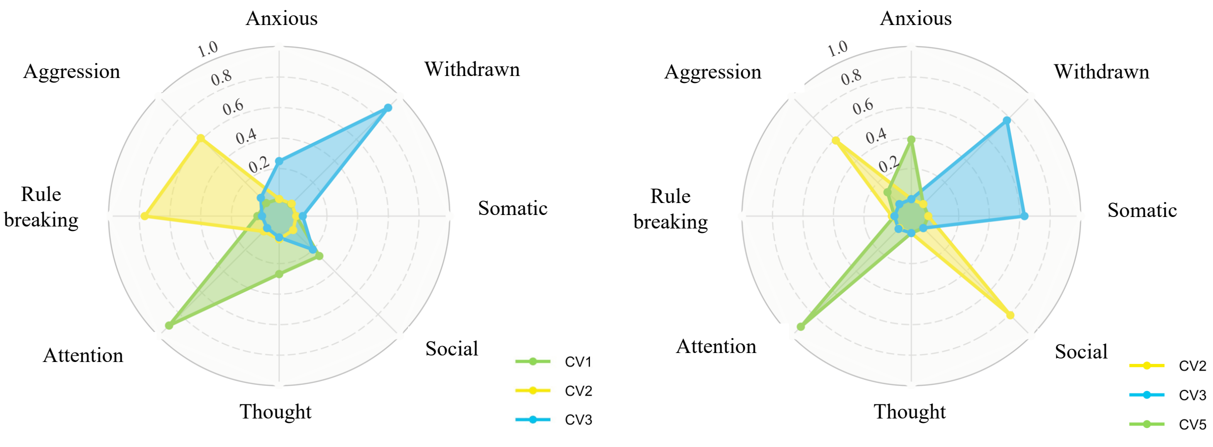


*Note.* The comparison of canonical loadings for CBCL syndrome scores in ABCD and Generation R. **a.** The canonical loadings of CBCL syndrome scores in ABCD. **b.** The canonical loadings of CBCL syndrome scores in Generation R. **CV1**: canonical variate 1, **CV2**: canonical variate 2, **CV3**: canonical variate 3.

**Supplementary Figure 2**

*Canonical loadings in Generation R in qualitative replication*

**a. CBCL loadings**

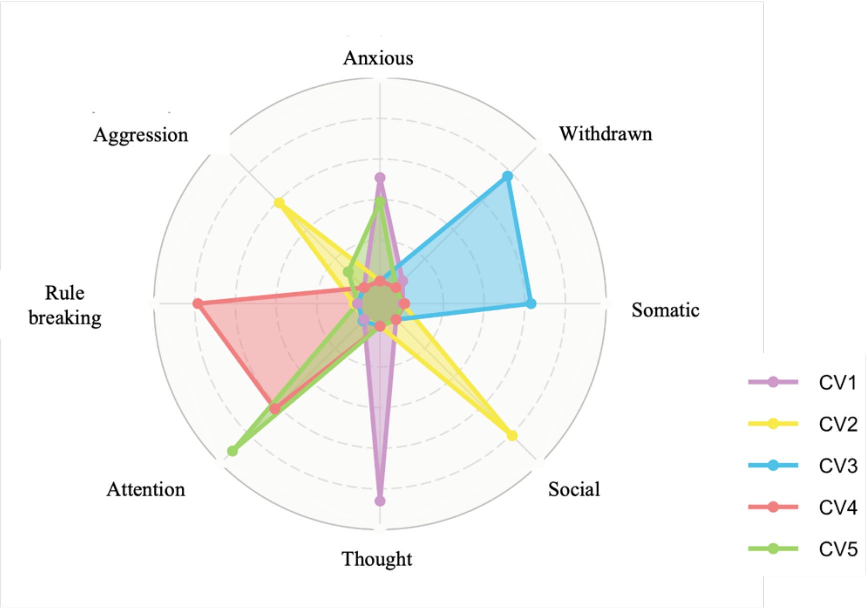


**b. Resting-state connectivity canonical variates**

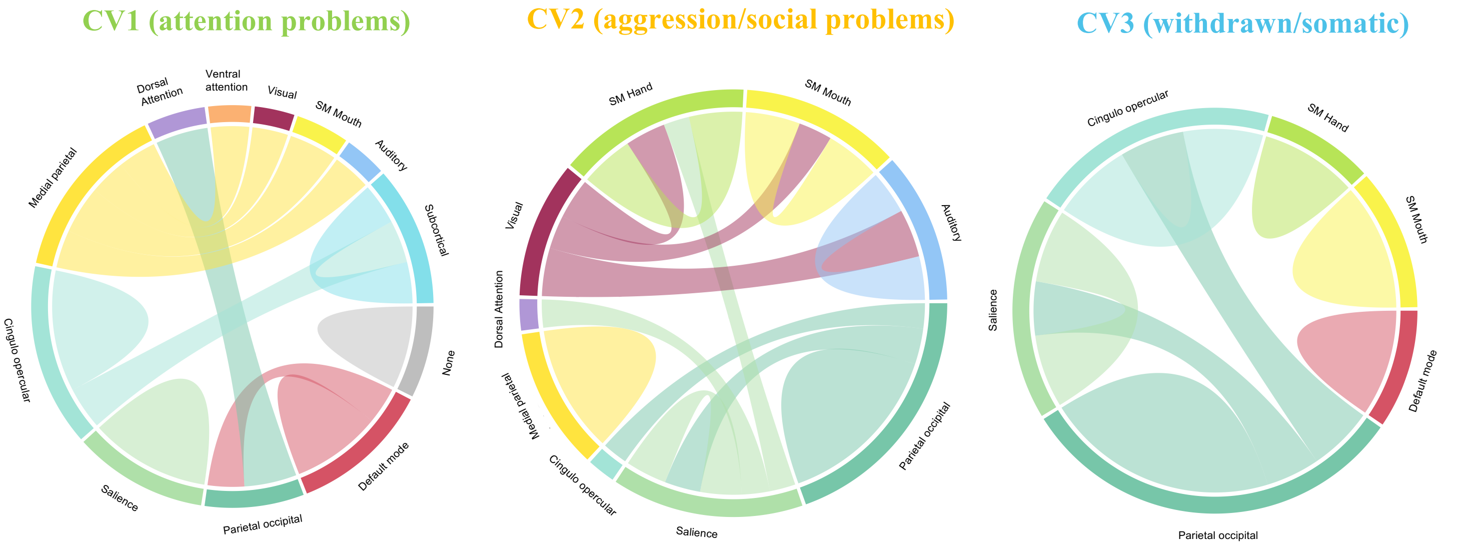


*Note.* ***a.*** *the five canonical variates survived permutation tests in Generation R.* **b.** brain connectivity modules involved in the three identified canonical variates in Generation R that are similar with ABCD.
